# Study protocol - Ascertaining the career Intentions of UK Medical Students (AIMS) post-graduation: a cross-sectional survey

**DOI:** 10.1101/2023.01.16.23284605

**Authors:** Tomas Ferreira, Alexander M. Collins, Rita Horvath

## Abstract

**Background:** Among doctors in the United Kingdom, there is growing sentiment regarding delaying specialist training, emigrating to practice medicine abroad, or leaving the profession altogether. This may have significant implications for the future of the profession in the UK. The extent to which this sentiment is also present in the medical student population is unknown.

**Methods:** The AIMS study is a national, multi-institution, cross-sectional study of all medical students at all medical schools in the United Kingdom. It will be administered via an online questionnaire and disseminated through a collaborative network recruited for this purpose. Our primary outcome is to determine current medical students’ career intentions after graduation and upon completing the Foundation Programme, and to establish the motivations behind these intentions. Secondary outcomes include determining which, if any, demographic factors alter the propensity to pursue different career paths available to a medical graduate, determining which specialties medical students plan on pursuing and understanding current views on the prospect of working in the National Health Service (NHS). Both quantitative analysis and thematic analysis will be used.

**Discussion:** Doctors’ career satisfaction within the NHS is a well-researched topic, however, there is a shortage of high-powered studies able to offer insight into medical students’ outlook on their future careers. It is anticipated that the results from this study will bring clarity to this issue. Identified areas of improvement in medical training or within the NHS could be targeted to improve doctors’ working conditions and help retain medical graduates. Results may also aid future workforce planning efforts.

**Trial Registration:** Not Applicable.

## Introduction/Background

In the United Kingdom, medical degrees typically last for 5 or 6 years, depending on whether one elects to pursue an additional year in the awarding of an intercalated degree. Alternatively, for previous degree holders, there is a 4-year accelerated medicine course. Presently, upon completion of their medical degree, graduates are provisionally registered with the General Medical Council (GMC). Graduates then enter the mandatory two-year UK Foundation Programme (UKFP), consisting of rotations in various settings and specialties. At the end of their first year of the UKFP, they are awarded full medical registration with the GMC. Doctors who have completed the UKFP are then able to pursue further training, which may be very specialised, for example, in ophthalmology or neurosurgery, or broader pathways such as in general practice or internal medicine training (1).

Historically, the vast majority of UKFP graduates pursued specialty training immediately after completing their foundation programme; for instance, in 2010, the proportion was 83.1% entering further training upon completing Foundation Year 2 (F2). However, the UK Foundation Programme Office (UKFPO) career destination report revealed that this number had fallen to only 34.9% of doctors in 2019 (2). The UKFPO has not repeated the survey since then, so it is difficult to surmise how these statistics may have changed in the interim.

In comparison to most other Organisation for Economic Co-operation and Development (OECD) countries, the UK has fewer doctors per population (3). One of the ways the British government has attempted to combat this issue is through the opening of new medical schools as well as increasing student capacity in existing medical schools (4, 5). It is our aim to understand the effectiveness of this approach; are the factors encouraging graduate doctors to seek alternatives to medicine in the UK, or in its entirety, also present in the undergraduate population?

AIMS also endeavours to investigate the reasons behind students’ intentions to both enter and leave the standard training pathway and attempt to identify any demographic and background information which might increase the propensity to leave training.

A BMA survey of 8,000 senior doctors determined that 44% of National Health Service (NHS) consultants in England plan to leave or take a break from working in the NHS over the next year (6). Similarly, a recent survey of 4,553 junior doctors in the NHS reported that 4 in 10 plan to leave the NHS, with 33% of these wanting to emigrate to another country to work (7). Broader literature supports these findings, with a 40-year survey series of junior doctors highlighting that the desire to remain in UK medical practice was lower in 2015 than had ever been recorded (8). Additionally, in 2021 nearly 10,000 doctors relinquished their licence to practice in the UK, making up nearly 1/10 of its total doctor workforce (5, 9). We aim to determine if this trend, found in both junior and senior doctors alike, may also manifest in the intentions of medical students yet to even enter the profession.

The study invites participants to share their views on many aspects of working in the NHS and how the prospect of working in the NHS could be improved. Data collected from this study could prove helpful in addressing some of the concerns of those making up the future of the profession. In Australia and New Zealand, a survey is distributed each year, where the Medical Deans of medical schools in the country survey final-year medical students and data is collected on their background, medical school experience, and any early geographical and specialty preferences for their future career (10). The information gained has had a lasting positive impact on curriculum development and future medical workforce planning. Additionally, it has assisted in workforce distribution and helped to plan specialist training pathways to meet the career interests of trainees and the public’s health needs.

The AIMS study is a national, multi-institution, cross-sectional study. AIMS endeavours to understand the factors involved in medical students’ decision-making around their future careers. Specifically, we are interested in students’ current career plans and why they may or may not intend to pursue specialty training, or a medical career, in the UK. We are also hoping to understand current views on the prospect of working in the NHS.

## Study Objectives

### Primary objectives

To determine current medical students’ career intentions after graduation and upon completing the Foundation Programme (FP), and to ascertain the motivations behind these intentions

### Secondary objectives

- To characterise the backgrounds of medical students intending to:

- emigrate to practice abroad
- leave medicine permanently
- undertake further study/career break
- not going into further medical training immediately after degree completion/foundation training
- To determine which specialties medical students plan on pursuing.
- To analyse medical students’ views on how the prospect of working in the NHS could be improved.

## Methods/Design

AIMS is a national, multi-centre, cross-sectional study of medical students. Participants’ responses will be recorded via an online survey platform. The survey contains 71 items; but no participants will view all items, it is dependent on the answers they give. The shortest question sequence involves 30 items, and the longest involves 43 items. Questions are structured using a combination of Likert scale matrices, multiple-choice options, and free-text entry to broaden the capture of sentiment nuance and improve precision in the data. Data collection will take place from the 16^th^ of January 2023, aiming to be completed on the 27^th^ of March 2023.

Part 1 of the survey involves a background and demographic section, which all participants will answer. Part 2 then focuses on career intentions immediately after graduation and after foundation training (for those entering). Part 3 invites participants to explore the factors behind their decision-making. Part 4 surveys which specialties the participants are currently inclined to pursue. The final part includes a free-entry text box where participants are invited to articulate how the prospect of working in the NHS could be improved and to obtain consent for follow-up.

The full 71 item questionnaire can be found in the Online Supplemental Materials.

To identify gaps in knowledge and inform the aim of the project, a review of existing literature was conducted, including similar questionnaires and qualitative studies on students’ perspectives. Feedback from senior clinical staff was also obtained. Questions were revised based on these reviewer comments to make them as non-directive as possible.

The survey will be hosted on the Qualtrics survey platform (Provo, Utah, USA), a GDPR-compliant online survey platform that supports mobile and desktop devices. The survey will be distributed through university mailing lists, student society pages, conferences, and personal and medical school social media platforms (such as Facebook, Instagram, LinkedIn, and Twitter). To maximize distribution across the UK, a national network of AIMS Collaborative members was recruited, representing all medical schools in the UK. Each member was asked to coordinate with their medical school dean to distribute the survey through official channels and regularly advertise it over the collection period from January to March.

A Participant Information Sheet (PIS) will be made available to all participants (available in the Online Supplemental Materials). This will explain the rationale, purpose, and voluntary nature of participation in the study. It will also emphasize that participation is anonymous, confidential, and voluntary and that participants can withdraw their consent at any time. Participants will be informed via the original advertisement leaflets and the front page of the survey that by submitting their answers, they consent for their data to be used. Email addresses will only be collected from participants who explicitly consent to this through the final question of the survey.

All data will be collected and stored on the secure online server Qualtrics. Survey data will be extracted from the software to a password-protected Microsoft Excel spreadsheet (Microsoft, California, USA) only available to the central study team. The University of Cambridge’s standards for data security will be followed for all data handling and record keeping.

### Study population

All current students of all years at UK medical schools recognized by the GMC and the Medical Schools Council are eligible to participate in the study. A list of eligible medical schools and approved programs can be found in the Online Supplemental Materials. It is worth noting that we have excluded schools that, although approved by the GMC, have not yet accepted their first cohort of students, and thus have no medical students to participate.

### Data collection

The survey will be disseminated primarily through recruited medical student collaborators recruited prior to the study launch. Collaborators will ensure that their medical school is formally engaged at an early stage of this study. They will be primarily responsible for disseminating this questionnaire among students at their medical school.

### Financial prize

At the end of data collection, a prize draw will be held for participants who have opted to provide their contact details. Two randomly selected participants will be selected at random to receive £50 in Amazon vouchers.

### Data Analysis

A sample size calculation was performed, and it was determined that a minimum of 8,026 participants are needed to have a confidence level of 95% that the results of the survey are within ±1% representation of the total medical student population. This calculation used a population size for UK medical students acquired via a Freedom of Information request to the GMC.

The STROBE cross-sectional study guidelines will be followed for this study (11). Descriptive statistics and comparisons between student year groups and other participant characteristics will be used. Appropriate statistical tests will be applied for these comparisons. For free-text questions from which qualitative data will be obtained, a thematic analysis will be carried out using the reflexive approach of Braun and Clarke (12).

### After Study Completion

Following study completion, teleconferences will be held with all collaborators to share and discuss the data analysis undertaken and the study results. Following this, the results will be presented at local, regional, national and international conferences. In addition, the results will be disseminated via publication in a peer-reviewed medical journal. All collaborators will be given PubMed-citable collaborative coauthorship under the institutional name ‘AIMS Collaborative’. We will have a hybrid authorship list of named authors and the institutional collaborative.

Medical school collaborators will be able to request their institution’s specific data and the analysis done on said data from the AIMS steering committee following study completion. This data will be anonymised. The fully anonymised data set will be made publicly available.

### Project timeline

Project timeline:
| Dates | Activity |
| --- | --- |
| 12 December 2022 - 9 January 2023 | Recruitment of collaborators throughout all UK medical schools via social media and communication with medical schools/societies. |
| 10 January 2023 - 15 January 2023 | Training of collaborators |
| 16 January 2023 - 27 March 2023 | Study runs nationally for 2 months |
| 28 March 2023 - 1 October 2023 | Data analysis and manuscript preparation |

This timeline will be approached with a level of flexibility, such as for centres with logistical obstacles to data collection.

## Discussion

While there have been: i) studies exploring which specialties junior doctors or medical students intend on pursuing (13, 14, 15, 16, 17, 18), ii) studies which focused on career intentions of those pursuing one specialty or exploring factors that attract them to one specific specialty (19, 20, 21, 22, 23, 24, 25, 26, 27, 28, 29, 30, 31, 32, 33, 34, 35), iii) studies specifically focused on reasons why doctors are leaving the UK (36, 37, 38, 39, 40), iv) studies exploring how medical students and junior doctors feel about specific aspects of working within the NHS (41, 42, 43, 44), v) studies investigating the desire for a career break post-foundation year 2 (also known as the ‘F3’) (45, 46, 47), there have been no recent, high-powered studies explicitly aimed at medical students, irrespective of current career ambitions or seniority, investigating overall career intentions and correlating it with demographic factors and medical student seniority. There has been one similar study, although focusing entirely on medical students and their intentions to leave the NHS, which is limited by its low power and lack of subanalysis regarding student seniority and demographic factor (48). We aim to address these limitations. Additionally, this study will be the first of a series of follow-up surveys of medical students, thus being the first medical students’ career intentions longitudinal study of its kind.

### Strengths

- Owing to its rigorous collaborator recruitment plans in hopes of high participant recruitment, this study will accurately demonstrate medical student intentions to either remain in the UK to practice or to go abroad as well as their specialty of choice – this could prove useful for workforce planning.
- The study will determine the current undergraduate consensus on several aspects of working in the NHS, which could help future policymakers prioritise change and better help medical schools prepare their junior students for the reality of such aspects.
- This study will determine which, if any, demographic factors alter the propensity to pursue different career paths available to a medical graduate.

### Limitations

- The study is limited by its snapshot nature, as we cannot see how career intentions have or will change over time. However, it is our intention to follow up with consenting participants and determine how their career intentions may have changed.
- It is possible that those already intending to exit the profession, to emigrate, or those disgruntled with the current circumstances will be more likely to complete the survey. To mitigate this, we will recruit over 150 collaborators who will distribute the survey in their respective medical schools, targeting all medical students. Additionally, the promotional material and the questions have been written not to imply any particular outcome is predicted or sought.
- It is possible that there might be challenges to the recruitment of collaborators or to the uptake of student responses which could jeopardise the study’s national goals.

This study presents a unique opportunity to accurately assess how medical students view their current career prospects and assess the feasibility of potentially implementing a regular, census-like survey of UK medical students to assist the government’s efforts in workforce planning and addressing grievances within the workforce, therefore resulting in higher job satisfaction.

## Supporting information

Participant Information Sheet

Ethics Approval Letter

AIMS Survey - Print

Eligible Medical Schools & Approved Programmes

## Data Availability

All data produced in the present study are available upon reasonable request to the authors

## Declarations

### Ethics approval and consent to participate

Ethical approval was granted by the University of Cambridge Research Ethics Committee (reference PRE.2022.124) on the 5th of January 2023.

### Consent for publication

Not applicable.

### Availability of data and materials

Not applicable.

### Competing interests

The authors declare that they have no competing interests.

### Funding

Queens’ College, University of Cambridge. The institution has had no role in the design of the study, and collection, analysis, and interpretation of data and in writing the manuscript.

### Authors’ contributions

TF responsible for conceptualisation. TF responsible for obtaining funding and ethical approval. TF responsible for drafting the manuscript. TF and AMC responsible for designing the survey. All authors responsible for editing and revising the manuscript. RH responsible for supervision. All authors have read and approved the manuscript.

## Acknowledgements

Not applicable.

## Authors’ information (optional)

Not applicable.

## Notes

### Competing Interest Statement

The authors have declared no competing interest.

### Funding Statement

This study was funded by Queens' College, University of Cambridge.

### Author Declarations

Ethics committee of the University of Cambridge gave ethical approval for this work

## Bibliography

1. How Doctor Specialty Training (Residency) Works in the UK. BMJ Careers. https://www.bmj.com/careers/article/how-doctor-specialty-training-residency-works-in-the-uk. Accessed 10 January 2023

2. UKFPO, UK Foundation Programme 2019 F2 Career Destinations Survey. 2019. Accessed 8 January 2023

3. Moberly T. UK has fewer doctors per person than most other OECD countries. BMJ: British Medical Journal. 2017;357.

4. Rimmer A. Five medical schools are created in England in bid to increase home grown doctors. BMJ: British Medical Journal (Online). 2018;360.

5. General Medical Council. The state of medical education and practice in the UK The workforce report 2022. 2022. Accessed 10 January 2023

6. BMJ Media Team. Catastrophic crisis facing NHS as nearly half of hospital consultants plan to leave in next year, warns BMA 2022. Available from: https://www.bma.org.uk/bma-media-centre/catastrophic-crisis-facing-nhs-as-nearly-half-of-hospital-consultants-plan-to-leave-in-next-year-warns-bma. Accessed 10 January 2023

7. Waters A. A third of junior doctors plan to leave NHS to work abroad in next 12 months. British Medical Journal Publishing Group; 2022.

8. Surman G, Goldacre MJ, Lambert TW. UK-trained junior doctors’ intentions to work in UK medicine: questionnaire surveys, three years after graduation. Journal of the Royal Society of Medicine. 2017;110(12):493–500.

9. NHS Workforce Statistics. December 2021 Doctors by Grade and Specialty. 2022.

10. Kaur B, Carberry A, Hogan N, Roberton D, Beilby J. The medical schools outcomes database project: Australian medical student characteristics. BMC Medical Education. 2014;14(1):1–10.

11. Cuschieri S. The STROBE guidelines. Saudi journal of anaesthesia. 2019;13(Suppl 1):S31.

12. Braun V, Clarke V. Using thematic analysis in psychology. Qualitative research in psychology. 2006;3(2):77–101.

13. Singh A, Alberti H. Why UK medical students change career preferences: an interview study. Perspectives on medical education. 2021;10(1):41–9.

14. Misky AT, Shah RJ, Fung CY, Sam AH, Meeran K, Kingsbury M, et al. Understanding concepts of generalism and specialism amongst medical students at a research-intensive London medical school. BMC Medical Education. 2022;22(1):1–11.

15. Lambert TW, Smith F, Goldacre MJ. Career specialty choices of UK medical graduates of 2015 compared with earlier cohorts: questionnaire surveys. Postgraduate Medical Journal. 2018;94(1110):191–7.

16. Surman G, Lambert TW, Goldacre MJ. Trends in junior doctors’ certainty about their career choice of eventual clinical specialty: UK surveys. Postgraduate medical journal. 2013;89(1057):632–7.

17. Svirko E, Goldacre MJ, Lambert T. Career choices of the United Kingdom medical graduates of 2005, 2008 and 2009: questionnaire surveys. Medical teacher. 2013;35(5):365–75.

18. Ibrahim M, Fanshawe A, Patel V, Goswami K, Chilvers G, Ting M, et al. What factors influence British medical students’ career intentions? Medical teacher. 2014;36(12):1064–72.

19. Reid K, Alberti H. Medical students’ perceptions of general practice as a career; a phenomenological study using socialisation theory. Education for Primary Care. 2018;29(4):208–14.

20. Rehman U, Sarwar MS, Brennan PA. Attitude of clinical medical students to Oral and Maxillofacial Surgery as a career: a perspective from two English Medical Schools. British Journal of Oral and Maxillofacial Surgery. 2022;60(4):448–53.

21. Barber S, Brettell R, Perera-Salazar R, Greenhalgh T, Harrington R. UK medical students’ attitudes towards their future careers and general practice: a cross-sectional survey and qualitative analysis of an Oxford cohort. BMC medical education. 2018;18(1):1–9.

22. Oliver H, Hudson B, Oliver C, Oliver M. UK undergraduate aspirations and attitudes survey: do we have a perception problem in clinical radiology? Clinical Radiology. 2020;75(2):158.e15-.e24.

23. Emmanouil B, Goldacre MJ, Lambert TW. Aspirations to become an anaesthetist: longitudinal study of historical trends and trajectories of UK-qualified doctors’ early career choices and of factors that have influenced their choices. BMC anesthesiology. 2017;17(1):1–9.

24. Barat A, Goldacre MJ, Lambert TW. Junior doctors’ early career choices do not predict career destination in neurology: 40 years of surveys of UK medical graduates. BMC medical education. 2019;19(1):1–9.

25. Robinson T, Lefroy J. How do medical students’ experiences inform their opinions of general practice and its potential as a future career choice? Education for Primary Care. 2022:1–9.

26. Tambyraja AL, McCrea CA, Parks RW, Garden OJ. Attitudes of medical students toward careers in general surgery. World journal of surgery. 2008;32(6):960–3.

27. Thomas A. What about forensic psychiatry as a career? Undergraduate and early post-graduate medical perspectives. Criminal Behaviour and Mental Health. 2012;22(4):247–51.

28. Smith F, Lambert TW, Pitcher A, Goldacre MJ. Career choices for cardiology: cohort studies of UK medical graduates. BMC medical education. 2013;13(1):1–8.

29. Maisonneuve JJ, Pulford C, Lambert TW, Goldacre MJ. Career choices for geriatric medicine: national surveys of graduates of 1974–2009 from all UK medical schools. Age and ageing. 2014;43(4):535–41.

30. Goodson AM, Payne KF, Tahim A, Cabot L, Fan K. Awareness of oral and maxillofacial surgery as a specialty and potential career pathway amongst UK medical undergraduates. The Surgeon. 2013;11(2):92–5.

31. Halder N, Hadjidemetriou C, Pearson R, Farooq K, Lydall GJ, Malik A, et al. Student career choice in psychiatry: findings from 18 UK medical schools. International Review of Psychiatry. 2013;25(4):438–44.

32. Goldacre MJ, Fazel S, Smith F, Lambert T. Choice and rejection of psychiatry as a career: surveys of UK medical graduates from 1974 to 2009. The British Journal of Psychiatry. 2013;202(3):228–34.

33. Pakpoor J, Handel AE, Disanto G, Davenport RJ, Giovannoni G, Ramagopalan SV. National survey of UK medical students on the perception of neurology. BMC medical education. 2014;14(1):1–5.

34. Sutton PA, Mason J, Vimalachandran D, McNally S. Attitudes, motivators, and barriers to a career in surgery: a national study of UK undergraduate medical students. Journal of surgical education. 2014;71(5):662–7.

35. Moore J, McDiarmid A, Johnston P, Cleland J. Identifying and exploring factors influencing career choice, recruitment and retention of anaesthesia trainees in the UK. Postgraduate medical journal. 2017;93(1096):61–6.

36. Wilson HC, Abrams S, Simpkin Begin A. Drexit: Understanding why junior doctors leave their training programs to train overseas: An observational study of UK physicians. Health Science Reports. 2021;4(4):e419.

37. Moss PJ, Lambert TW, Goldacre MJ, Lee P. Reasons for considering leaving UK medicine: questionnaire study of junior doctors’ comments. BMJ. 2004;329(7477):1263.

38. Sharma A, Lambert TW, Goldacre MJ. Why UK-trained doctors leave the UK: cross-sectional survey of doctors in New Zealand. Journal of the royal society of medicine. 2012;105(1):25–34.

39. Milner A, Nielsen R, Verdery AM. Brexit and the European National Health Service England Workforce: A Quantitative Analysis of Doctors’ Perceived Professional Impact and Intentions to Leave the United Kingdom. Annals of global health. 2021;87(1).

40. Lambert TW, Smith F, Goldacre MJ. Why doctors consider leaving UK medicine: qualitative analysis of comments from questionnaire surveys three years after graduation. Journal of the Royal Society of Medicine. 2018;111(1):18–30.

41. Scanlan GM, Cleland J, Johnston P, Walker K, Krucien N, Skåtun D. What factors are critical to attracting NHS foundation doctors into specialty or core training? A discrete choice experiment. BMJ open. 2018;8(3):e019911.

42. Ryan C, Ward E, Jones M. Recruitment and retention of trainee physicians: a retrospective analysis of the motivations and influences on career choice of trainee physicians. QJM: An International Journal of Medicine. 2018;111(5):313–8.

43. Lachish S, Goldacre MJ, Lambert T. Associations between perceived institutional support, job enjoyment, and intentions to work in the United Kingdom: national questionnaire survey of first year doctors. BMC medical education. 2016;16(1):1–8.

44. Cleland JA, Johnston P, Watson V, Krucien N, Skåtun D. What do UK medical students value most in their careers? A discrete choice experiment. Medical Education. 2017;51(8):839–51.

45. Church HR, Agius SJ. The F3 phenomenon: Early-career training breaks in medical training. A scoping review. Medical Education. 2021;55(9):1033–46.

46. Hollis AC, Streeter J, Van Hamel C, Milburn L, Alberti H. The new cultural norm: reasons why UK foundation doctors are choosing not to go straight into speciality training. BMC Medical Education. 2020;20(1):1–9.

47. Agius SJ, Tack G, Murphy P, Holmes S, Hayden J. Why do medical trainees take time out of their specialty training programmes? British Journal of Hospital Medicine. 2014;75(10):584–9.

48. Cutting J, Adil Salim Elbakri S, Adams H, Jaunoo S. A national survey of medical student career ambitions. The Bulletin of the Royal College of Surgeons of England. 2020;102(1):44–7.

