## Supplementary material for "Study protocol - Ascertaining the career Intentions of UK Medical Students (AIMS) post-graduation: a cross-sectional survey": Participant Information Sheet

**What is the aim of this study?** This study aims to determine current medical students' career intentions post-graduation and post-foundation training, to identify factors involved in decision making for students' career choices and to analyse medical students' views on how the prospect of working in the NHS could be improved.

**Why have I been selected to take part?** All medical students currently studying at UK medical schools recognised by the General Medical Council (GMC) are being invited to take part in the questionnaire.

**What do I have to do?** If you decide to participate in this study, you will be asked to complete a questionnaire about your background, your career intentions after graduation and after foundation training, and your motivations for these answers. This study is voluntary. If you choose to participate, you will be asked to complete the survey by clicking on the link found at the end of this document. This survey is expected to take about 4-7 minutes to complete, but there is no time limit. No background knowledge is required. By submitting the survey, you consent to the collection and storage of data in accordance with the UK General Data Protection Regulation (GDPR) within the survey. For more information on GDPR please click on the following link: https://gdpr-info.eu.

**Do I have to participate?** Participation is entirely voluntary. You may withdraw at any point during the questionnaire for any reason, before submitting your answers, by closing the browser. In cases of withdrawal from the study prior to submission of the survey, no data is recorded. If you have already submitted data and wish to withdraw from the study, please contact by 31st March 2023.

**Who has approved this study**? This study has been reviewed and approved by the University of Cambridge's Research Ethics Committee on the 5th of January 2023, reference PRE.2022.124.

**How will my data be used?** All answers will be anonymous, and we will take all reasonable precautions to ensure that they remain confidential. Data will be stored in a password-protected file and may be used in academic publications. Your IP address will not be stored. After completion of data collection, no email addresses will be stored unless you consent to being followed up via the survey's final question. Prior to completion of data collection, we will store your institutional email address to confirm your student status. Research data will be stored for a minimum of ten years after publication or public release.

**Who will have access to my data?** Qualtrics is the data controller of the personal data held about you and, as such, will determine how your personal data are used. Their privacy notice can be found here: https://www.qualtrics.com/privacy-statement. Qualtrics will share any email address you provide and your anonymised responses with the University of Cambridge, for the purposes of research as highlighted above. Researchers involved in the project will have access to this anonymised data.

**Are there any benefits to taking part?** Although there are no immediate individual benefits to participating in this survey, you are given the opportunity to contribute to research which may impact you. You may find this survey an opportunity to self-reflect on your career plans after you graduate. Additionally, all participants will be entered into a prize draw for one of two £50 Amazon vouchers.

**Will the research be published?** The findings of this study may be published in peer-reviewed journals, presented at conferences and a summary of the findings will be made available on social media.

AIMS UNIVERSITY OF CAMBRIDGE School of Clinical Medicine

**Are there any possible risks involved with my participation?** There are no anticipated disadvantages, side effects, risks, and/or discomforts of taking part in this study. If participating in the study leads to distress, you may stop the survey at any time. If your distress continues after leaving the survey, we have provided a list of supportive services nationwide that can be helpful and that you might consider contacting (appears at the close of survey).

**Who do I contact if I have a concern about the study or I wish to complain?** If you have a concern about any aspect of this project, please speak to the principal researcher. If you remain unhappy or wish to make a formal complaint, please contact the Research and Information Governance, School of Clinical Medicine, University of Cambridge:.

**How do I find out what was learned in this study?** This study is expected to be completed by April 2023. If you would like a brief summary of the results, please write to us by email to request information

**Link to the survey:** https://cambridge.eu.qualtrics.com/jfe/form/SV_cx55RTspDLTIzWK

Kind Regards,

**Tomas Ferreira**
AIMS Study Lead

**Dr. Rita Horvath**
Supervisor, Director of Research, Horvath Laboratory, Department of Clinical Neurosciences, University of Cambridge
