## Supplementary material for "Study protocol - Ascertaining the career Intentions of UK Medical Students (AIMS) post-graduation: a cross-sectional survey": Ethics Approval Letter

*Milly Bodfish*
*Secretary*

Dr Rita Horvath
Department of Clinical Neuroscience
University of Cambridge

UNIVERSITY OF CAMBRIDGE

CAMBRIDGE
PSYCHOLOGY RESEARCH
ETHICS COMMITTEE

5 January 2023

Application No: PRE.2022.124

Dear Dr Horvath

**Ascertaining the career Intentions of UK Medical Students' post-graduation: a cross-sectional survey**

The Cambridge Psychology Research Ethics Committee has given ethical approval to your research project *Ascertaining the career Intentions of UK Medical Students' post-graduation: a cross-sectional survey* as set out in your application submitted on 8 December 2022.

The Committee attaches certain standard conditions to all ethical approvals. These are:

(a) that if the staff conducting the research should change, any new staff should read the application submitted to the Committee for ethical approval and this letter (and any subsequent letter concerning this application for ethical approval). An amendment request should also be submitted (guidance at www.bio.cam.ac.uk/psyres/amendments);
(b) that if the procedures used in the research project should change or the project itself should be changed, you should consider whether it is necessary to submit a further application for any modified or additional procedures to be approved;
(c) that if the employment or departmental affiliation of the staff should change, you should notify us of that fact.

Members of the Committee also ask that you inform them should you encounter any unexpected ethical issues.

Ethical approval will expire 31 August 2023. If you require an extension, please submit an amendment request before the expiry date (guidance here: https://www.bio.cam.ac.uk/psyres/amendments).

If you would let us know that you are able to accept these conditions, we will record that you have been given ethical approval.

Yours sincerely

M Bodfish
