## Supplementary material for "Study protocol - Ascertaining the career Intentions of UK Medical Students (AIMS) post-graduation: a cross-sectional survey": AIMS Survey - Print

### Demographics

Thank you for taking part in the study. Please note that participating in this survey is entirely optional.

In 2010, 83.1% of Foundation Year 2 (F2) doctors went into further training. In 2018, this number was only 37.7%. This represents a significant change in the makeup of doctors in the UK on a backdrop of a wider NHS staffing challenge. AIMS endeavours to understand the factors involved in medical students' decision-making around their future career. Specifically, we are interested in what students' current career plans are, and why they may, or may not choose to pursue specialty training, or a medical career more broadly, in the UK. We are also hoping to understand current views on the prospect of working in the NHS.

Ethical approval was granted by the University of Cambridge Research Ethics Committee (PRE.2022.124) on 5 January 2023.

All responses will remain confidential. Your email address will only be visible to the study leads and will be deleted from our records once all data has been collected (unless you consent to being followed up at the end of the survey) and there is no need for further communication.

You may withdraw from the study at any point by contacting.

By submitting your answers to the survey, you consent to us collecting this data and acknowledging that anonymised data may be published and used for purposes beyond this study.

All participants will be entered into a prize draw for one of two £50 Amazon vouchers.

**I understand that my participation is voluntary and that I am free to withdraw at any time without giving a reason and I consent to participate in this study.**

○ Yes

**Email Address**

Please enter your institutional email address (ending in 'ac.uk'. We will use this to verify your student status and we may contact you to notify you of a prize win or for clarification of responses). Please ensure there are no spaces at the end of your email.

**Age**

**Gender**

**Which of the below options best describes your ethnicity?**

#### University

#### Year of study - Please read description.

(as of September 2022)

- If you are in your fourth year of study and it is your final year, please select final year (i.e., GEM)

- If you are in your fourth year of study but it is your penultimate year, please select penultimate year.

- If you are currently intercalating, please select your current year of study (e.g., intercalating between 3rd and 4th year on a 5 year course please select Year 4).

- ○ Year 1
- ○ Year 2
- ○ Year 3
- ○ Year 4 (not penultimate year)
- ○ Penultimate year
- ○ Final year

#### What is your expected graduation year?

#### Do you have a previous or intercalated degree?

- ○ Yes, prior to studying Medicine.
- ○ Yes, an intercalated degree.
- ○ Yes, both.
- ○ Not yet, but intend on intercalating.
- ○ Not yet, but currently intercalating.
- ○ No.

#### What is your student fee status?

- ○ Home
- ○ EU
- ○ International (Non-EU)

**Did you, at any point in your education, attend a fee-paying independent school?**

E.g., private school.

- ○ Yes
- ○ No
- ○ Prefer not to say

#### Intentions

**Do you intend to join the NHS Foundation Programme after graduation?**

- ○ Yes - plan to complete F1 & F2
- ○ Yes - plan to complete F1 & emigrate to practice abroad
- ○ Yes - plan to complete F1 & leave medicine permanently.
- ○ No - plan on leaving medicine permanently.
- ○ No - plan on emigrating
- ○ No - plan on taking a break or undertaking further study.

**What do you intend to do after completing the NHS Foundation Programme?**

- ○ Enter specialty training in the UK
- ○ Non-training clinical job in the UK, e.g. 'F3 year', JCF or CTF
- ○ Emigrating to practice medicine abroad (including temporarily)
- ○ Taking a break or undertaking further study
- ○ Leaving medicine permanently

**You have indicated your intention to leave medicine permanently. In which industry do you plan to work after leaving medicine? If unsure, please enter N/A"**

#### In which country do you intend to practice?

If you are unsure, please enter N/A.

#### Reasons for emigrating to practice abroad

In your previous answers, you have indicated your intentions to practice medicine abroad.

Please indicate the level of importance of the below factors in your decision making

| | Very important | Important | Neither important nor unimportant | Not important | Not at all important |
|---|---|---|---|---|---|
| Remuneration or pay at junior level | ◯ | ◯ | ◯ | ◯ | ◯ |
| Remuneration or pay at consultant level | ◯ | ◯ | ◯ | ◯ | ◯ |
| Work-life balance | ◯ | ◯ | ◯ | ◯ | ◯ |
| Family | ◯ | ◯ | ◯ | ◯ | ◯ |
| Desire for a life change | ◯ | ◯ | ◯ | ◯ | ◯ |
| Ease of entry into training (competition) | ◯ | ◯ | ◯ | ◯ | ◯ |
| Length of training | ◯ | ◯ | ◯ | ◯ | ◯ |
| Standard of training | ◯ | ◯ | ◯ | ◯ | ◯ |
| Ability to choose work location | ◯ | ◯ | ◯ | ◯ | ◯ |
| Working conditions of a doctor in the NHS | ◯ | ◯ | ◯ | ◯ | ◯ |
| Uncertainty about which specialty to pursue | ◯ | ◯ | ◯ | ◯ | ◯ |

#### You have indicated that you intend to emigrate to practice medicine, do

#### you intend on returning to the UK?

- ◯ Yes - after a few years
- ◯ Yes - after I complete my training
- ◯ No

#### Reasons for leaving medicine permanently

In your previous answers, you have indicated your intentions to leave medicine permanently.

Please indicate the level of importance of the below factors in your decision making.

| | Very important | Important | Neither important nor unimportant | Not important | Not at all important |
|---|---|---|---|---|---|
| Remuneration or pay at junior level | ◯ | ◯ | ◯ | ◯ | ◯ |
| Remuneration or pay at consultant level | ◯ | ◯ | ◯ | ◯ | ◯ |
| Work-life balance | ◯ | ◯ | ◯ | ◯ | ◯ |
| Family | ◯ | ◯ | ◯ | ◯ | ◯ |
| Desire for a life change | ◯ | ◯ | ◯ | ◯ | ◯ |
| Ability to choose work location | ◯ | ◯ | ◯ | ◯ | ◯ |
| Working conditions of a doctor in the NHS | ◯ | ◯ | ◯ | ◯ | ◯ |
| Stress levels associated with profession | ◯ | ◯ | ◯ | ◯ | ◯ |
| Burnout | ◯ | ◯ | ◯ | ◯ | ◯ |

#### Reasons for not entering specialty training immediately after F2

In your previous answers, you have indicated your intentions to not enter specialty training immediately after completing your F2 year. Please indicate the level of importance of the below factors in your decision making.

| | Very | | Neither important nor | Not | Not at all |
|---|---|---|---|---|---|

| | important | Important | unimportant | important | important |
|---|---|---|---|---|---|
| Saving money or greater earning potential. | | | | | |
| Undertaking further study | | | | | |
| Portfolio building for applications | | | | | |
| Travel | | | | | |
| Experiencing another healthcare system | | | | | |
| Starting a family | | | | | |
| Reapplying for specialty programme. | | | | | |
| Ability to choose work location | | | | | |
| Burnout | | | | | |
| Uncertainty about which specialty to pursue | | | | | |

#### Reasons for not entering foundation training immediately after graduation

In your previous answers, you have indicated your intentions to not enter foundation training immediately after graduation. Please indicate the level of importance of the below factors in your decision making.

| | Very important | Important | Neither important nor unimportant | Not important | Not at all important |
|---|---|---|---|---|---|
| Undertaking further study | | | | | |
| Travel | | | | | |
| Experiencing another healthcare system | | | | | |
| Starting a family | | | | | |

| | | | | | |
|---|---|---|---|---|---|
| Reapplying for specialty programme. | ○ | ○ | ○ | ○ | ○ |
| Ability to choose work location | ○ | ○ | ○ | ○ | ○ |
| Health | ○ | ○ | ○ | ○ | ○ |
| Burnout | ○ | ○ | ○ | ○ | ○ |

#### Views on a career in the NHS

For each of the points below, how would you describe your level of satisfaction regarding their current status in the NHS?

| | Very satisfied | Satisfied | Neither satisfied nor unsatisfied | Not satisfied | Not at all satisfied |
|---|---|---|---|---|---|
| Remuneration or pay at junior level | ○ | ○ | ○ | ○ | ○ |
| Remuneration or pay at consultant level | ○ | ○ | ○ | ○ | ○ |
| Work-life balance | ○ | ○ | ○ | ○ | ○ |
| Ability to choose work location | ○ | ○ | ○ | ○ | ○ |
| Ease of entry into training (competition) | ○ | ○ | ○ | ○ | ○ |
| Length of training | ○ | ○ | ○ | ○ | ○ |
| Standard of training | ○ | ○ | ○ | ○ | ○ |
| Working conditions of a doctor in the NHS | ○ | ○ | ○ | ○ | ○ |
| Exposure to desired specialty during foundation programme | ○ | ○ | ○ | ○ | ○ |
| Theatre time during foundation programme | ○ | ○ | ○ | ○ | ○ |
| Cost of training (i.e., mandatory exams, courses, memberships) | ○ | ○ | ○ | ○ | ○ |
| Length of time to decide on a specialty | ○ | ○ | ○ | ○ | ○ |
| Pension Tax rules as a consultant | ○ | ○ | ○ | ○ | ○ |

| | | | | | |
|---|---|---|---|---|---|
| Overall satisfaction with the prospect of working in the NHS | ○ | ○ | ○ | ○ | ○ |

**Are you certain about which specialty you wish to pursue?**

- ○ Very certain
- ○ Somewhat certain
- ○ Neither certain nor uncertain
- ○ Somewhat uncertain
- ○ Very uncertain

**Which specialty (or specialties) most interest you?**

Select **up to** a maximum of 3 options (if you are certain, please select only one)

- ☐ Acute internal medicine
- ☐ Allergy
- ☐ Anaesthetics
- ☐ Audio vestibular medicine
- ☐ Cardio-thoracic surgery
- ☐ Cardiology
- ☐ Clinical genetics
- ☐ Clinical neurophysiology
- ☐ Clinical oncology
- ☐ Community sexual and reproductive health
- ☐ Dermatology
- ☐ Emergency medicine
- ☐ Endocrinology and diabetes mellitus
- ☐ Gastro-enterology
- ☐ General practice
- ☐ General surgery
- ☐ Genito-urinary medicine
- ☐ Geriatric medicine
- ☐ Haematology
- ☐ Histopathology
- ☐ Immunology

- [ ] Infectious diseases
- [ ] Intensive care medicine
- [ ] Medical microbiology
- [ ] Medical oncology
- [ ] Neurology
- [ ] Neurosurgery
- [ ] Nuclear medicine
- [ ] Obstetrics and gynaecology
- [ ] Occupational medicine
- [ ] Ophthalmology
- [ ] Oral and maxillo-facial surgery
- [ ] Otolaryngology (ENT)
- [ ] Paediatric surgery
- [ ] Paediatrics
- [ ] Palliative medicine
- [ ] Pathology
- [ ] Plastic surgery
- [ ] Psychiatry
- [ ] Public health medicine
- [ ] Radiology
- [ ] Rehabilitation medicine
- [ ] Renal medicine
- [ ] Respiratory medicine
- [ ] Rheumatology
- [ ] Sport and exercise medicine
- [ ] Trauma and orthopaedic surgery
- [ ] Tropical medicine
- [ ] Urology
- [ ] Vascular surgery

**What steps could be taken to improve the prospect of working in the NHS?**

(Optional)

**Do you consent to being contacted by us for potential follow-up studies regarding your career intentions?**

We will store your email address to contact you in the future.

- ◯ Yes
- ◯ No

Powered by Qualtrics
