## Supplementary material for "Study protocol - Ascertaining the career Intentions of UK Medical Students (AIMS) post-graduation: a cross-sectional survey": Eligible Medical Schools & Approved Programmes

Eligible Medical Schools and Approved Programmes

A combination of the universities of Dundee and St. Andrews (ScotGEM)

A combination of the University of Brighton and the University of Sussex

A combination of the University of Hull and the University of York

Anglia Ruskin School of Medicine

Aston Medical School

Brunel University London Medical School

Cardiff University

Edge Hill University Medical School

Imperial College London

Keele University

Kent and Medway Medical School

King's College London

Lancaster University

Queen Mary University of London

St George's University of London

Swansea University

The Queen's University of Belfast

The University of Aberdeen

The University of Birmingham

The University of Bristol

The University of Buckingham

The University of Cambridge

The University of Central Lancashire

The University of Dundee

The University of Dundee

The University of East Anglia

The University of Edinburgh

The University of Exeter

The University of Glasgow

The University of Leeds

The University of Leicester

The University of Liverpool

The University of Manchester

The University of Newcastle

The University of Nottingham

The University of Oxford

The University of Plymouth

The University of Sheffield

The University of Southampton

The University of St Andrew's

The University of Warwick

Ulster University School of Medicine

University College London

University of Chester Medical School

University of Sunderland School of Medicine

Excluded for lack of cohort at time of recruitment:

- University of Chester Medical School
- Three Counties Medical School
